# The Pattern of Metformin Discontinuation and Reinitiation in US adults with Prediabetes

**DOI:** 10.64898/2025.11.29.25341250

**Authors:** Zeshui Yu, Chen Jiang, Zheng Zeng, Hui Wang, Yuqing Chen, Lirong Wang

## Abstract

**Background:** The American Diabetes Association recommends that metformin may be considered for patients with prediabetes. However, evidence on discontinuation and reinitiation patterns of metformin use in patients with prediabetes remains limited

**Objective:** To describe the rates and specific patient characteristics associated with metformin discontinuation and reinitiation in patients with prediabetes.

**Method:** We conducted a retrospective observational study using longitudinal electronic health record data from the Truveta database. We identified a total of 23,911 new metformin users with a baseline A1C between 5.7% and less than 6.5% in an incident cohort of prediabetes from January 1, 2019, to May 31, 2025. The pattern of metformin utilization was calculated using linked medication dispensing records within the Truveta network. Treatment discontinuation and reinitiation following the first discontinuation were estimated using the Kaplan–Meier model. Cox proportional hazards regression models were applied to evaluate the association between patient characteristics and treatment discontinuation.

**Result:** In this retrospective cohort, 14,857 patients (62.13%) discontinued metformin during a maximum follow-up of 6 years. Compared with those who continued treatment, patients who discontinued were more likely to be female (10,137 [68.23%] vs 5,642 [62.31%]), have baseline A1C <6% (5,455[36.72%] vs 2,892 [31.94%]), 60 years or younger (5,190 [57.32%] vs 5,190 [57.32%] ), have baseline class 3 obesity (4,434 [26.7%] vs 2,029 [20.6%]). The median time to discontinuation was 0.82 years (95% CI, 0.79–0.85), with 27.15% of patients discontinuing in 90 days after initiation. The cumulative proportion of discontinuation at 1 year and 2 years was 54.45% (95% CI, 53.77–55.13%) and 69.48% (95% CI, 68.77–70.91%), respectively. The proportion of patients reinitiating metformin after the first discontinuation at 1 year and 2 years was 32.44% (95% CI, 31.64–33.25%) and 41.81% (95% CI, 40.89–42.75%), respectively. The median time to reinitiation was 3.81 years (95% CI, 3.44–4.08)

**Conclusion:** Most patients with prediabetes discontinued metformin within the first year, and reinitiation was limited, highlighting gaps in adherence and the need for strategies to optimize metformin use in this population.

## Introduction

An estimated one in three adults in the United States has prediabetes, representing a major public health challenge and contributing to substantial healthcare expenditures.^1–3^ Lifestyle modification, including dietary changes and increased physical activity, remains the cornerstone of diabetes prevention.^4–6^ However, despite the high prevalence and clinical burden of prediabetes, no pharmacological treatment has been formally approved by the US Food and Drug Administration (FDA) for the prevention of type 2 diabetes mellitus (T2DM).^7,8^

Metformin, a widely used and cost-effective generic medication, has been proposed as an alternative or adjunctive off-label intervention for individuals with prediabetes.^9^ It has been associated with improvements in glycemic control and insulin sensitivity.^10,11^ Due to its favorable tolerability and safety profile and affordability, metformin continues to be considered as a preferred treatment for diabetes prevention in selecting high-risk populations among other antidiabetic medications. Although the American Diabetes Association (ADA) stated the treatment of metformin can be considered in patients with prediabetes, previous studies have reported that only 0.1% to 4% of patients with prediabetes receive the prescription for metformin, reflecting the low prevalence of prescribing metformin in diabetes prevention.^12–21^ The reasons for the low utilization of metformin among individuals with prediabetes are not fully understood but have been postulated to include inconsistencies among clinical guidelines, limited evidence regarding its long-term efficacy in preventing T2DM and its macrovascular and microvascular complications.^8,22^

Several randomized clinical trials have demonstrated the safety and efficacy of metformin in reducing the incidence of diabetes compared to placebos in patients with prediabetes.^23,24^ Despite its potential benefits, little is known about the long-term patterns of metformin utilization among patients with prediabetes, including discontinuation and subsequent reinitiation. It is important to examine metformin use patterns in patients with prediabetes, as suboptimal adherence and persistence could diminish its clinical benefits observed in clinical trials^25^ The ADA recommends considering metformin therapy for individuals under 60 years of age with a BMI > 35 kg/m² or an HbA1c > 6.0%, as these high-risk groups are more likely to experience preventive benefits from metformin.^12,23^ In this high-risk population, premature discontinuation or intermittent use of metformin may significantly diminish its preventive benefits and undermine the clinical outcomes, representing a major barrier to effective treatment.^25,26^

To address these gaps, we aimed to assess the rate of metformin discontinuation and reinitiation and identify clinical factors associated with the discontinuation of the metformin in a large incident cohort of patients with prediabetes using real-world electronic health record (EHR) data.

## Methods

### Study design

This retrospective observational study was conducted in accordance with the Strengthening the Reporting of Observational Studies in Epidemiology (STROBE) guidelines. Patient consent was waived due to the use of de-identified data.^23^ The data analysis was performed from May 2025 to July 2025.

### Data source

Truveta is an expanding health data platform that aggregates continuously updated, linked, de-identified structured EHR data from a network of 30 U.S. healthcare systems, representing approximately 120M patient population across the United States. Truveta compiles longitudinal, real-world EHR data, including demographics, diagnoses, medications, procedures, laboratory results, and vital signs. In addition to capturing clinical care delivered within Truveta-affiliated health systems, the dataset is additionally supplemented with pharmacy dispensing data obtained through external linkages, providing a more comprehensive view of patient behavior and real-world medication use. Medication dispensing data includes details such as fill dates, days’ supply, and quantities dispensed.^27,28^ Collectively, these features enhance the completeness of longitudinal medical histories, enabling efficient examination of real-world medication utilization patterns and outcomes using EHR data.

### Patient population

First, we identified an incident cohort of patients receiving the first diagnosis of prediabetes between January 1, 2019, and December 31, 2024, from the Truveta EHR database. Patients were required to have at least two diagnosis codes for prediabetes, identified using ICD-10 or SNOMED terminologies. To ensure complete baseline information and improve observability, patients were required to meet the following criteria: (1) have at least one year of previous medical history, defined as ≥1 valid encounter occurring at least 365 days before the first prediabetes diagnosis code; and (2) demonstrate regular engagement with the Truveta Health System, defined as ≥1 valid encounter within 1 to 365 days before the first prediabetes diagnosis code. In addition, patients were required to have at least one follow-up encounter after the date of the prediabetes diagnosis. Patients were excluded if they met criteria for type 2 diabetes prior to the date of first prediabetes diagnosis, defined as the presence of ≥1 diagnosis code for type 2 diabetes, evidence of insulin or DPP-4 inhibitor use, or HbA1c >6.5%.^27^ Individuals with any documented diagnosis of type 1 diabetes or gestational diabetes were also excluded.^27^

In the prediabetes cohort, patients were included if they initiated metformin between 7 days before and any time after their first prediabetes diagnosis, to account for potential delays in recording the diagnosis. To ensure adequate follow-up and outcome measurement, we restricted analyses to patients with at least one follow-up encounter and a recorded baseline HbA1c measurement between 5.7% and <6.5% within the window of 90 days before to 7 days after metformin initiation. Patients with an A1C value <5.7% or >= 6.5% before or within 7 days after metformin initiation were excluded to ensure the cohort represented individuals without normoglycemia or established type 2 diabetes. Patients who developed type 2 diabetes, type 1 diabetes, or gestational diabetes before metformin initiation were excluded. Additionally, patients who had only a single dispensation with days of supply shorter than 30 days were excluded. All data obtained from the Truveta database were deidentified in accordance with the Health Insurance Portability and Accountability (HIPAA) Privacy Rule. Therefore, this study was exempt from institutional review board approval.

### Measures

Metformin dispensations were identified using RxNorm codes recorded in the medication dispensation records. Discontinuation was defined as a gap of ≥90 days without any metformin on hand. In the Truvata database, this was calculated by taking the interval between two consecutive fill dates and subtracting the estimated days of medication on hand, which included leftover pills from prior fills plus the days of supply from the current prescription. Intervals exceeding 90 days were classified as discontinuation. Changes in dosage were not considered discontinuation if the subsequent dispensation occurred within 90 days. To account for excess supply from prior fills, the dispensing date, leftover pills, and the days of supply were used to estimate medication availability. Summary statistics showed that a 90-day supply was the most common maximum prescription length for metformin in both the study database and national U.S. prescribing patterns.^29^ Therefore, we selected a ≥90-day gap without a dispensation to capture clinically meaningful treatment interruptions rather than shorter breaks. Reinitiation, assessed descriptively as a secondary outcome, was defined as the first subsequent dispensing of metformin after primary discontinuation.

### Statistical analysis

Descriptive statistics were performed among patients at baseline, stratified by those with and without discontinuation during the follow-up using two-sample t-tests and chi-square tests as appropriate. Both discontinuation and reinitiation were treated as time-to-event outcomes and were subject to censoring. In discontinuation analyses, the date when patients received their first metformin dispensing was defined as the index time (time 0). Patients were followed until the earliest of the following events: the first occurrence of discontinuation, the last encounter, the study end date (May 31st, 2025), or a maximum follow-up duration of six years. To determine the time to the first metformin discontinuation, Kaplan–Meier curves were utilized to estimate the proportion of patients remaining on the metformin treatment at 1,2,3,4,5 and 6 years after initiation. Patients who discontinued metformin were included in the reinitiation analysis. The date of their first discontinuation was defined as the index date. A similar Kaplan-Meier analysis was also performed to estimate the time to the first reinitiation among those who discontinued metformin. The cumulative risk and proportion of at-risk patients who discontinued and subsequently reinitiated metformin were calculated from each model.

Multivariable adjusted Cox proportional regression models were built to assess the risk of time to the first discontinuation of metformin. The proportional hazards assumption was tested using Schoenfeld residuals with both global and covariate-specific tests at p<0.05. Violations were addressed by stratification or inclusion of time-by-covariate interaction terms. Covariates were selected based on previous studies examining metformin adherence and persistence or offering the descriptive statistic of the prevalence of metformin among patients with prediabetes. The model-building process involved two stages. First, univariate Cox regression models were fitted for each related covariate. All covariates with a univariate p-value of <0.10 were then eligible for inclusion in the multivariable model. A stepwise selection procedure (with entry and stay criteria set at p<0.05) was used to identify the final set of independent risk factors from this pool. The final multivariable model included demographic covariates (age, gender, race/ethnicity, income), condition factor (time from prediabetes diagnosis), physiological factors (baseline BMI and A1C), and comorbidities (cardiovascular disease, chronic kidney disease, depression and anxiety, heart failure, hyperlipidemia, hypertension, hypothyroidism, and nonalcoholic steatohepatitis).

## Results

### Baseline demographics and clinical characteristics

Among the 23,911 patients initiating metformin for prediabetes, 14,857 (62.1%) discontinued treatment during a median follow-up of 0.95 years. Compared with patients who continued metformin, those who discontinued were younger (mean age 52.0 vs. 55.9 years), more likely to be female (68.2% vs. 62.3%), and Black (17.5% vs. 10.2%,). They also had higher baseline BMI (36.9 vs. 35.1 kg/m²), lower baseline A1C (6.04% vs. 6.07%,), and shorter median time from prediabetes diagnosis to metformin initiation (0.58 vs. 0.98 years, all p<0.001; Table 1).

**Table 1.** Baseline Demographics and Clinical Characteristics of Prediabetes Patients Initiating Metformin.

|  |  | Patients, No. (%) |  |  |  |  |
| --- | --- | --- | --- | --- | --- | --- |
|  |  | Missing | Overall | Continued | Discontinued | P-Value |
| n |  |  | 23911 | 9054 | 14857 |  |
| Age<60 |  |  | 15449 (64.61) | 5190 (57.32) | 10259 (69.05) | <0.001 |
| Age>=60 |  |  | 8462 (35.39) | 3864 (42.68) | 4598 (30.95) |  |
| Age, y |  |  | 53.45 (14.54) | 55.91 (14.16) | 51.95 (14.56) | <0.001 |
| Sex | Female |  | 15779 (65.99) | 5642 (62.31) | 10137 (68.23) | <0.001 |
|  | Male |  | 8132 (34.01) | 3412 (37.69) | 4720 (31.77) |  |
| Race | White |  | 15643 (65.42) | 6187 (68.33) | 9456 (63.65) | <0.001 |
|  | Asian |  | 1767 (7.39) | 827 (9.13) | 940 (6.33) |  |
|  | Black |  | 3518 (14.71) | 920 (10.16) | 2598 (17.49) |  |
|  | Other or unknown |  | 2983 (12.48) | 1120 (12.37) | 1863 (12.54) |  |
| Ethnicity | Hispanic or Latino |  | 3422 (14.31) | 1132 (12.50) | 2290 (15.41) | <0.001 |
|  | Not Hispanic or Latino |  | 19418 (81.21) | 7479 (82.60) | 11939 (80.36) |  |
|  | Unknown |  | 1071 (4.48) | 443 (4.89) | 628 (4.23) |  |
| Individual income | <=30000 |  | 3837 (16.05) | 1216 (13.43) | 2621 (17.64) | <0.001 |
|  | 30001-50000 |  | 1123 (4.70) | 380 (4.20) | 743 (5.00) |  |
|  | 50001-80000 |  | 4268 (17.85) | 1578 (17.43) | 2690 (18.11) |  |
|  | >80001 |  | 3708 (15.51) | 1722 (19.02) | 1986 (13.37) |  |
|  | Unknown |  | 10975 (45.90) | 4158 (45.92) | 6817 (45.88) |  |
| Marital status | Married or partnered |  | 12848 (53.73) | 5136 (56.73) | 7712 (51.91) | <0.001 |
|  | Unmarried |  | 7804 (32.64) | 2645 (29.21) | 5159 (34.72) |  |
|  | Other |  | 3259 (13.63) | 1273 (14.06) | 1986 (13.37) |  |
| Baseline BMI class | BMI<35 |  | 9778 (40.89) | 4204 (46.43) | 5574 (37.52) | <0.001 |
| | BMI≥35 | | 10175 (42.55) | 3457 (38.18) | 6718 (45.22) | |
|  | Unknown |  | 3958 (16.55) | 1393 (15.39) | 2565 (17.26) |  |
| Baseline BMI, kg/m<sup>2</sup> |  | 3958 | 36.24 (8.37) | 35.13 (8.20) | 36.93 (8.40) | <0.001 |
| Time from prediabetes, y |  |  | 0.73 (1.12) | 0.98 (1.31) | 0.58 (0.96) | <0.001 |
| A1C, % |  |  | 6.05 (0.22) | 6.07 (0.22) | 6.04 (0.22) | <0.001 |
| A1C < 6.0% |  |  | 8347 (34.91) | 2892 (31.94) | 5455 (36.72) | <0.001 |
| A1C ≥ 6.0% | | | 15564 (65.09) | 6162 (68.06) | 9402 (63.28) | |
| Cardiovascular disease |  |  | 2998 (12.54) | 1331 (14.70) | 1667 (11.22) | <0.001 |
| Chronic kidney disease |  |  | 1324 (5.54) | 555 (6.13) | 769 (5.18) | 0.002 |
| Coronary artery disease |  |  | 2011 (8.41) | 928 (10.25) | 1083 (7.29) | <0.001 |
| Cerebrovascular disease |  |  | 1251 (5.23) | 529 (5.84) | 722 (4.86) | 0.001 |
| Depression and anxiety |  |  | 12162 (50.86) | 4502 (49.72) | 7660 (51.56) | 0.006 |
| Heart failure |  |  | 758 (3.17) | 294 (3.25) | 464 (3.12) | 0.622 |
| Hyperlipidemia |  |  | 16719 (69.92) | 6971 (76.99) | 9748 (65.61) | <0.001 |
| Hypertension |  |  | 13789 (57.67) | 5471 (60.43) | 8318 (55.99) | <0.001 |
| Hypothyroidism |  |  | 4599 (19.23) | 1809 (19.98) | 2790 (18.78) | 0.023 |
| Nonalcoholic steatohepatitis |  |  | 646 (2.70) | 244 (2.69) | 402 (2.71) | 0.993 |
| Peripheral artery disease |  |  | 199 (0.83) | 96 (1.06) | 103 (0.69) | 0.003 |
| Follow-up, y |  |  | 0.95 [0.25, 1.24] | 1.41 [0.52, 1.95] | 0.66 [0.25, 0.83] | <0.001 |

**Abbreviations:** n, number; y, year; BMI, body mass index; kg, kilogram; m, meter; A1C, hemoglobin A1C.
T-tests were used to compare continuous variables, and chi-square tests were applied to categorical variables. Variables were considered significantly associated with metformin discontinuation if the p-value was <0.05. Clinical conditions were identified using ICD codes. Cardiovascular disease was defined as the presence of coronary artery disease, cerebrovascular disease, or peripheral artery disease.

The discontinuation group also had lower prevalence of most comorbidities, including cardiovascular disease (11.2% vs. 14.7%), coronary artery disease (7.3% vs. 10.3%), heart failure (3.1% vs. 3.3%), hyperlipidemia (65.6% vs. 77.0%), hypertension (56.0% vs. 60.4%), and chronic kidney disease (5.2% vs. 6.1%) (all p ≤ 0.05 except heart failure and nonalcoholic steatohepatitis, which did not differ significantly).

Among patients who discontinued metformin, those who reinitiated treatment (n =5,535) were younger (mean age 51.74 vs. 53.13 years, *p*<0.001), more likely to be male (35.28% vs. 29.68%, *p*<0.001), and of Black (18.28% vs. 17.01%, *p*=0.002) or Asian (7.05% vs. 5.90%, *p*=0.002) race/ethnicity compared with those who did not reinitiate (**Supplementary Table 2)**.

### Metformin Dispensing Pattern

**Table 2** summarizes metformin dispensing patterns. Overall, 5,535 patients had at least two treatment periods, with most experiencing two periods (n = 3,990), 1,100 having three, and 445 having four or more. Following the first discontinuation, the median time to reinitiation was 150 days (IQR, 90–304 days), reflecting a wide range of interruption duration. Among patients who reinitiated metformin, the median duration of the secondary treatment period was 180 days (IQR, 90–361 days).

**Table 2.** Descriptive Statistics for Metformin Dispensing Pattern.

| Measurements | Statistic | Values |
| --- | --- | --- |
| Number of metformin treatment periods* |  |  |
| Twice | n | 3,990 |
| Three times | n | 1,100 |
| ≥ 4 times | n | 445 |
| Time from the first discontinuation to the first reinitiation (days) | n | 5,535 |
|  | Mean (SD) | 242.19 |
|  | 25th percentile | 90 |
|  | Median (50th percentile) | 150 |
|  | 75th percentile | 304 |
| Time from the first reinitiation to censor (days)** | Mean (SD) | 279.32<br>(302.84) |
|  | 25th percentile | 90 |
|  | Median (50th percentile) | 180 |
|  | 75th percentile | 361 |

\*A treatment period was defined as a continuous duration of metformin use without a gap of more than 90 days without medication on hand.
\*A new treatment period was considered to begin if a patient reinitiated metformin after a gap of >90 days without metformin available.
\*\*Censoring was defined as the second occurrence of metformin discontinuation after a gap of >90 days without metformin available, the last clinical encounter before the study end date (August 11, 2025), or reaching a maximum follow-up duration of six years, whichever came first.

### Proportion of discontinuation and reinitiation

Kaplan-Meier analysis demonstrated a high and rapid rate of metformin discontinuation (**Figure 1**). Out of 23,911 patients, 14,857 (62.7%) discontinued metformin during a maximum follow-up of 6 years. The median time to discontinuation was 0.85 years (95% CI: 0.79–0.85), with a substantial proportion of patients discontinuing within the first 90 days (27.15%). Over half of the patients had discontinued the medication one year after initiation (54.45%, 95% CI: 53.77–55.31%), with the rate increasing to 69.48% (95% CI: 68.77–70.19%) by the second year. Among those who discontinued, the proportion of patients who reinitiated metformin after the first discontinuation was 32.44% (95% CI: 31.64–33.25%) at 1 year and 41.81% (95% CI: 40.89–42.75%) at 2 years, respectively. The median time to reinitiation was 3.81 years (95% CI: 3.44–4.08), and just over half of the patients (58.4%) had reinitiated metformin by 6 years after their primary discontinuation

**Figure 1.**
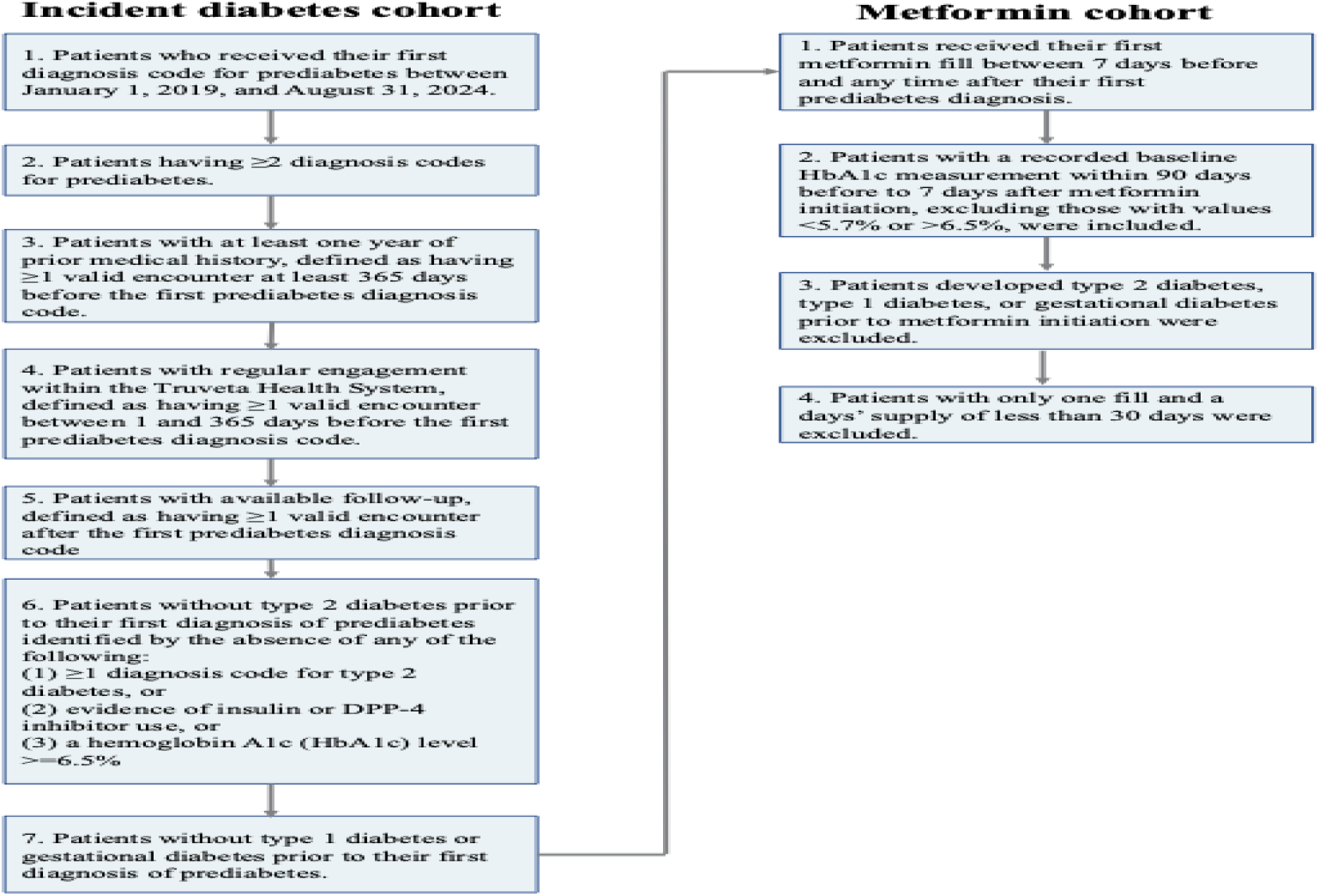
Inclusion Criteria.

**Figure 2.**
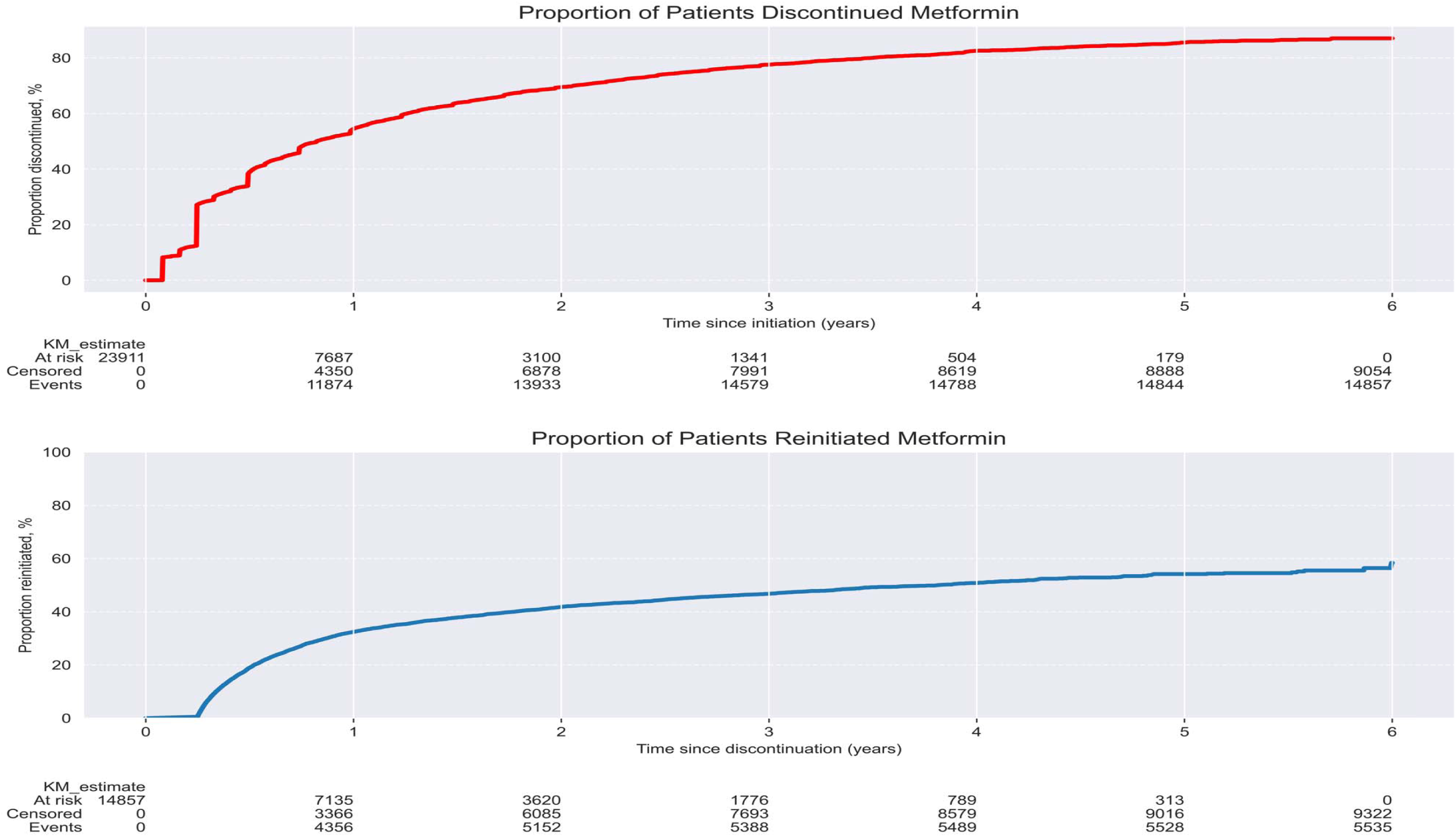
Cumulative Discontinuation and Reinitiation Rates of Metformin Among Patients With Prediabetes. In discontinuation analysis, patients were followed until the earliest of the following events: the first occurrence of the discontinuation, the last encounter, the study end date (May 31st, 2025), or a maximum follow-up duration of six years in discontinuation analysis. In reintiation analysis, patients were followed until the earliest of the following events: the first occurrence of the reinitiation, the last encounter, the study end date (May 31st, 2025), or a maximum follow-up duration of six years.

**Figure 3.**
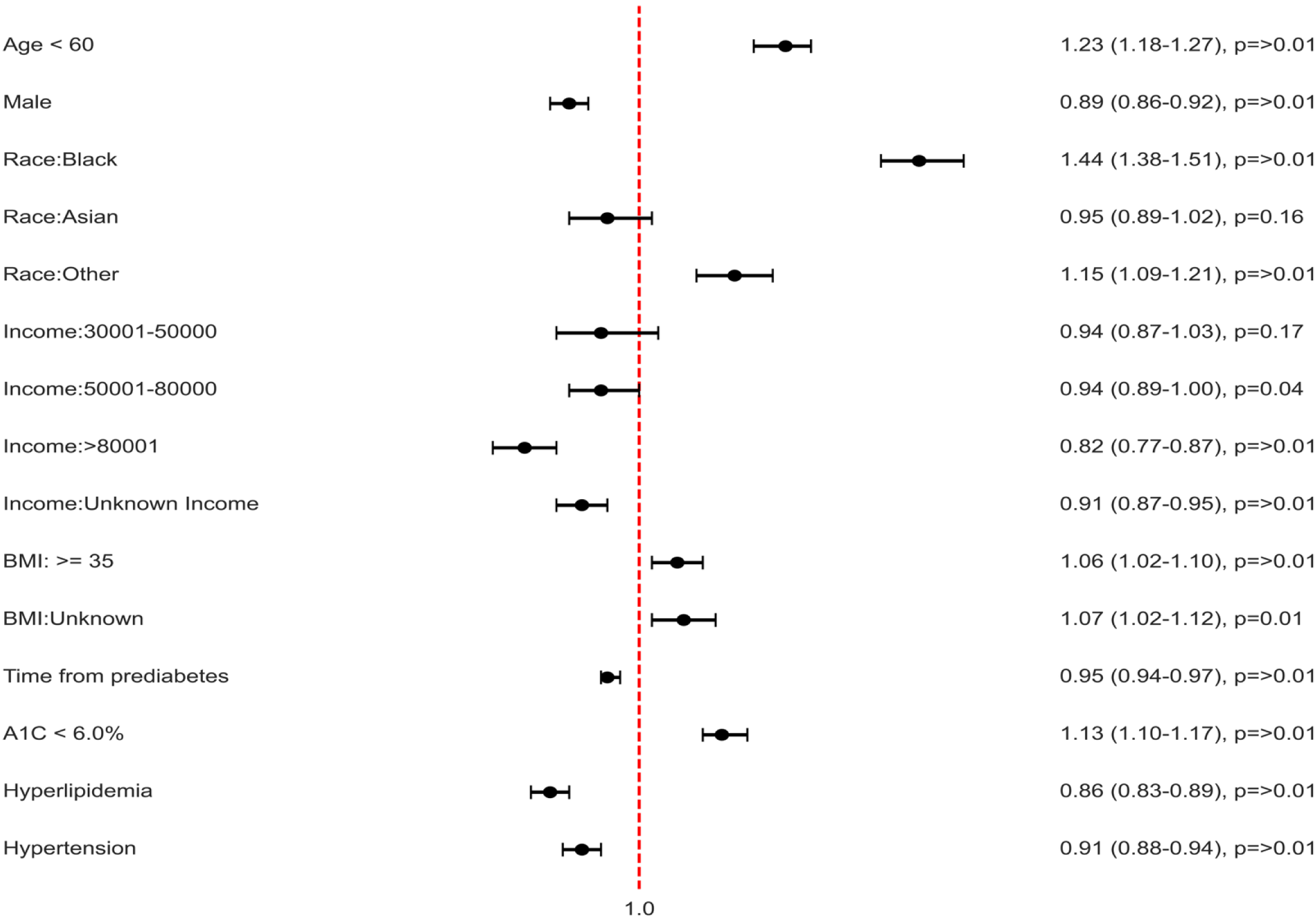
Forest Plot of Associations Between Patient Characteristics and Discontinuation. Abbreviations: BMI, Body mass index; Hemoglobin A1C, This multivariable Cox proportional hazards model was constructed by including independent risk factors identified through univariate analysis, with variable selection performed via a stepwise algorithm.

### Multivariable Analysis of Discontinuation Risk

In the multivariable Cox regression analysis (**Table 3**), patients aged <60 years had a 23% higher hazard of discontinuation compared to those ≥60 years (Adjusted Hazard Ratio [aHR]: 1.23, 95% CI: 1.18–1.27). Black patients had a 44% higher hazard compared to White patients (aHR: 1.44, 95% CI: 1.38–1.51). Higher baseline BMI (≥35 kg/m² vs. <35 kg/m², aHR: 1.06, 95% CI: 1.02–1.10) and baseline A1C <6.0% (aHR: 1.13, 95% CI: 1.10–1.17) were also associated with a greater risk of discontinuation. Conversely, male sex (aHR: 0.89, 95% CI: 0.86–0.92) and higher income levels (e.g., >$80,001 vs. ≤$30,000, aHR: 0.82, 95% CI: 0.77–0.87) were associated with a lower discontinuation risk. A longer duration between prediabetes diagnosis and metformin initiation was associated with lower discontinuation risk (aHR per unit increase: 0.95, 95% CI: 0.94–0.97). In addition, hyperlipidemia (aHR: 0.86, 95% CI: 0.83–0.89) and hypertension (aHR: 0.91, 95% CI: 0.88–0.94) were independently associated with a lower hazard of discontinuing metformin.

**Table 3.** Cumulative Discontinuation and Reinitiation Rate of Metformin in Patients with Prediabetes.

| Time | Proportion of patients discontinued metformin (CI) | Proportion of patients reinitiated metformin (CI) |
| --- | --- | --- |
| Baseline | 0.00% (0.00% - 0.00%) | 0.00% (0.00% - 0.00%) |
| 1 month | 8.26% (7.91% - 8.61%) | 0.00% (0.00% - 0.00%) |
| 2 months | 10.89% (10.50% - 11.29%) | 0.00% (0.00% - 0.00%) |
| 3 months | 27.15% (26.58% - 27.72%) | 0.00% (0.00% - 0.00%) |
| 0.5 years | 38.82% (38.30% - 39.57%) | 18.89% (18.32% - 19.61%) |
| 1 year | 54.45% (53.77% - 55.13%) | 32.44% (31.64% - 33.25%) |
| 1.5 years | 63.80% (63.13% - 64.53%) | 37.82% (36.98% - 38.71%) |
| 2 years | 69.48% (68.77% - 70.19%) | 41.81% (40.89% - 42.75%) |
| 2.5 years | 74.09% (73.36% - 74.83%) | 44.57% (43.59% - 45.56%) |
| 3 years | 77.54% (76.78% - 78.29%) | 46.74% (45.70% - 47.79%) |
| 3.5 years | 80.00% (79.20% - 80.78%) | 49.21% (48.07% - 50.37%) |
| 4 years | 82.54% (81.68% - 83.39%) | 50.79% (49.56% - 52.04%) |
| 4.5 years | 84.02% (83.13% - 84.98%) | 52.86% (51.45% - 54.27%) |
| 5 years | 85.49% (84.45% - 86.50%) | 54.17% (52.61% - 55.74%) |
| 5.5 years | 86.47% (85.32% - 87.57%) | 54.52% (52.90% - 56.16%) |
| 6 years | 87.02% (85.75% - 88.24%) | 58.39% (53.90% - 62.94%) |

Abbreviation: CI, confidence interval.
The proportion of patients who discontinued and subsequently reinitiated metformin was estimated using the Kaplan–Meier estimator.

**Table 4.**
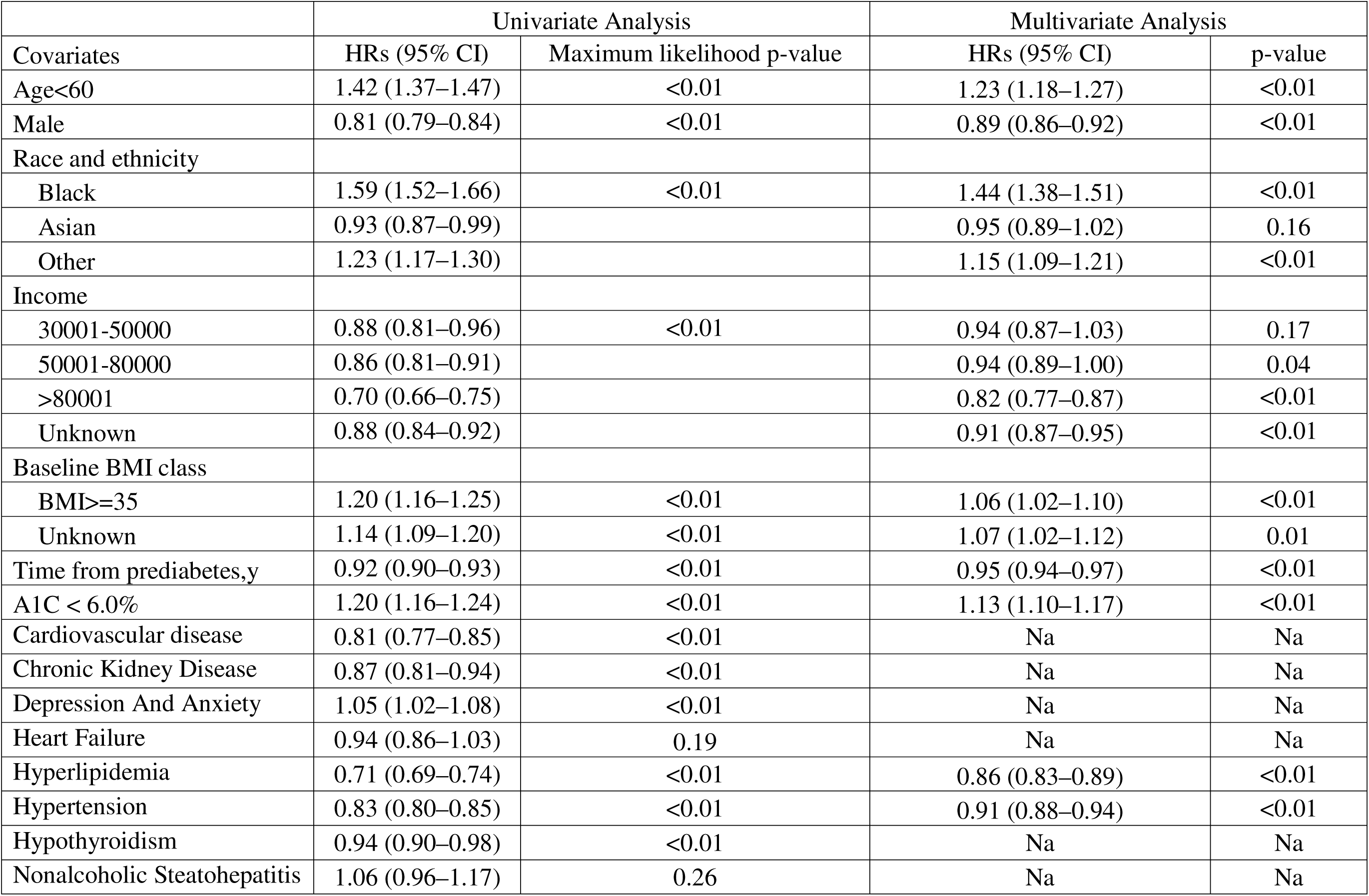
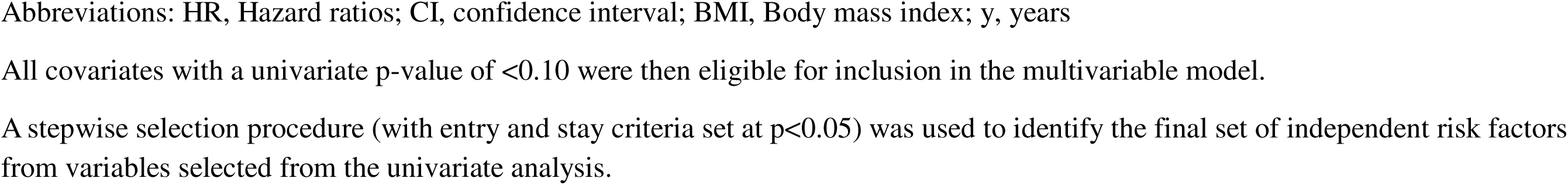
Associations Between Covariates and Discontinuation Outcomes for Patients with Prediabetes.

|  | Univariate Analysis |  | Multivariate Analysis |  |
| --- | --- | --- | --- | --- |
| Covariates | HRs (95% CI) | Maximum likelihood p-value | HRs (95% CI) | p-value |
| Age<60 | 1.42 (1.37–1.47) | <0.01 | 1.23 (1.18–1.27) | <0.01 |
| Male | 0.81 (0.79–0.84) | <0.01 | 0.89 (0.86–0.92) | <0.01 |
| Race and ethnicity |  |  |  |  |
| Black | 1.59 (1.52–1.66) | <0.01 | 1.44 (1.38–1.51) | <0.01 |
| Asian | 0.93 (0.87–0.99) |  | 0.95 (0.89–1.02) | 0.16 |
| Other | 1.23 (1.17–1.30) |  | 1.15 (1.09–1.21) | <0.01 |
| Income |  |  |  |  |
| 30001-50000 | 0.88 (0.81–0.96) | <0.01 | 0.94 (0.87–1.03) | 0.17 |
| 50001-80000 | 0.86 (0.81–0.91) |  | 0.94 (0.89–1.00) | 0.04 |
| >80001 | 0.70 (0.66–0.75) |  | 0.82 (0.77–0.87) | <0.01 |
| Unknown | 0.88 (0.84–0.92) |  | 0.91 (0.87–0.95) | <0.01 |
| Baseline BMI class |  |  |  |  |
| BMI≥35 | 1.20 (1.16–1.25) | <0.01 | 1.06 (1.02–1.10) | <0.01 |
| Unknown | 1.14 (1.09–1.20) | <0.01 | 1.07 (1.02–1.12) | 0.01 |
| Time from prediabetes,y | 0.92 (0.90–0.93) | <0.01 | 0.95 (0.94–0.97) | <0.01 |
| A1C < 6.0% | 1.20 (1.16–1.24) | <0.01 | 1.13 (1.10–1.17) | <0.01 |
| Cardiovascular disease | 0.81 (0.77–0.85) | <0.01 | Na | Na |
| Chronic Kidney Disease | 0.87 (0.81–0.94) | <0.01 | Na | Na |
| Depression And Anxiety | 1.05 (1.02–1.08) | <0.01 | Na | Na |
| Heart Failure | 0.94 (0.86–1.03) | 0.19 | Na | Na |
| Hyperlipidemia | 0.71 (0.69–0.74) | <0.01 | 0.86 (0.83–0.89) | <0.01 |
| Hypertension | 0.83 (0.80–0.85) | <0.01 | 0.91 (0.88–0.94) | <0.01 |
| Hypothyroidism | 0.94 (0.90–0.98) | <0.01 | Na | Na |
| Nonalcoholic Steatohepatitis | 1.06 (0.96–1.17) | 0.26 | Na | Na |

Abbreviations: HR, Hazard ratios; CI, confidence interval; BMI, Body mass index; y, years
All covariates with a univariate p-value of $<0.10$ were then eligible for inclusion in the multivariable model.
A stepwise selection procedure (with entry and stay criteria set at $p<0.05$ ) was used to identify the final set of independent risk factors from variables selected from the univariate analysis.

## Discussion

In this comprehensive analysis of metformin discontinuation and reinitiation patterns among patients with prediabetes over 6 years, we provide valuable real-world insights into medication utilization patterns and behaviors in this at-risk population. We identified most individuals discontinued the metformin within the first year, with a significant proportion of individuals who discontinued the treatment in less than or equal to 90 days. Reinitiation rates following initial discontinuation were also alarmingly low. Additionally, we identified several factors associated with early discontinuation of metformin, including female sex, Black race, age under 60 years, BMI ≥35 kg/m², and A1C <6.0%. These findings highlight the suboptimal long-term persistence to metformin in this population and have practical implications for developing more tailored strategies to improve treatment persistence.

Several randomized clinical trials have reported that the pharmacological treatment of metformin was associated with a lower incidence of diabetes among individuals with prediabetes.^24,30^ Actual adherence and persistence may differ from those observed in clinical trials, thereby attenuating the treatment effect. Our findings show that persistence with metformin was suboptimal, given that a persistence rate of ≥80% is generally considered optimal.^25,31^ This is particularly concerning given metformin’s role as a foundational therapy in prediabetes. Premature discontinuation or intermittent use of metformin may undermine its preventive potential and represent a significant barrier to effective treatment in prediabetes.

Additionally, non-persistence of metformin use may also contribute to variability in blood glucose and body weight, which are known risk factors for adverse cardiovascular outcomes.^32,33^

Several factors may be postulated to explain the high discontinuation rates and low reinitiation rates observed in this study. First, metformin has not been formally approved by the FDA for prediabetes. This regulatory uncertainty may contribute to hesitancy around long-term metformin use among both clinicians and patients. Second, adverse effects, including gastrointestinal intolerance and vitamin B12 deficiency, may prompt patients to discontinue therapy or favor lifestyle interventions over pharmacologic treatment.^8^ Third, concerns about the cost-effectiveness and potential overtreatment of metformin in prediabetes may lead to metformin discontinuation, given that about one-third of patients with prediabetes revert to normoglycemia and two-thirds will not progress to T2DM without treatment.^34^ Uncertainty about the metformin’s ability to alter disease pathophysiology and its unclear long-term benefits may further reduce motivation to continue therapy.^13^ Finally, the growing availability of alternative agents, such as GLP-1 receptor agonists, has gained prominence for delaying progression to type 2 diabetes and providing additional cardiovascular and renal benefits. This may influence clinical decisions to switch the pharmacotherapy among patients who are likely to benefit from these newer therapies.

According to the recommendation by the American Diabetes Association, metformin may be considered for specific subgroups of patients with prediabetes, including individuals under 60 years of age, those with a BMI ≥35, women with a history of gestational diabetes, or patients with elevated fasting plasma glucose (≥110 mg/dL) or HbA1c levels (≥6.0%).^12^ Our findings indicate that patients under 60 years of age are significantly more likely to discontinue metformin treatment compared to those aged 60 or older, suggesting that age is an important factor for metformin persistence. Patients in young adulthood and midlife represent a promising opportunity for pharmacological intervention in prediabetes, as metformin is more effective in preventing T2DM in those under 60 years of age compared to individuals aged 60 years or older. However, the generally higher rates of medication non-adherence among younger adults compared to older adults present unique barriers and specific needs within this population.^35^ The high discontinuation rate may be attributed to several patient-related factors, including limited engagement with the healthcare system, competing care and family priorities, and lack of prioritization on long-term health. Additionally, this age group falls below the Medicare eligibility threshold. Although metformin is generally considered an affordable medication, our findings show a strong association between income and treatment discontinuation. It is imperative that the medical community and policymakers implement tailored approaches to address the preferences and needs of this population, with the goal of improving metformin adherence and persistence.

Patients with a baseline A1C ≥6% were more likely to persist with metformin, consistent with the current recommendation by current guidelines for those with elevated HbA1c. However, patients with a baseline BMI of 35 or higher were associated with higher rates of metformin discontinuation compared to those with lower BMI. Ensuring persistence in metformin use among this population is particularly important, as metformin has been shown to be as effective as lifestyle modification in preventing type 2 diabetes among individuals with a BMI ≥ 35.^30^ Given that patients with obesity often face challenges in maintaining lifestyle interventions, metformin represents a valuable alternative strategy, and promoting persistence with therapy is crucial to fully realize its preventive benefits, including T2DM risk reduction and potential weight loss. Baseline hyperlipidemia and hypertension were associated with better adherence and persistence, potentially because these patients have more regular interactions with the healthcare system due to the chronic nature of their conditions.^36,37^

This study has several strengths. To our best knowledge, this is the first study to comprehensively evaluate the rate of discontinuation and subsequent reinitiation patterns of metformin in patients with prediabetes. Previous observational studies have mainly reported low metformin prescribing rates (1.1%–4%) using cross-sectional designs, with a slight upward trend over time.^12–21^ Using survival design with longitudinal data, our findings extend current evidence on metformin use by providing novel insights into metformin utilization patterns, including persistence measured by abrupt discontinuation or long dispensing gaps, and the subsequent reinitiation of therapy in patients with prediabetes. Additionally, the use of linked EHR data with pharmacy dispensing records allowed us to include patients paying for metformin out of pocket and capture their weight and baseline A1C at the initiation of medication.

Our study should be interpreted in the context of several limitations. First, we were unable to determine the specific reasons for medication discontinuation. For instance, gastrointestinal intolerance is the most common adverse effect of metformin and may contribute to discontinuation; however, such adverse events are often difficult to capture from EHR. Second, lifestyle data were not available, despite lifestyle modification remaining the cornerstone for preventing progression to diabetes. Third, our cohort included only patients with formal clinical diagnoses of prediabetes. Those with impaired fasting glucose, impaired glucose tolerance, or elevated HbA1c levels without receiving any formal diagnosis code were not enrolled. Restricting the cohort to ICD-coded cases allowed us to focus on patients formally recognized by clinicians as having prediabetes and potentially eligible for intervention. Finally, we were unable to identify women with a history of gestational diabetes, which may influence metformin persistence.

## Conclusion

Despite strong evidence and guideline recommendations supporting metformin use in prediabetes management, our drug utilization analysis indicates that successful management of prediabetes may be undermined by suboptimal treatment persistence. Premature metformin discontinuation could contribute to poorer clinical outcomes and a greater overall disease burden. A more systematic and tailored approach is needed to identify high-risk patients and implement targeted strategies to improve metformin persistence in this population.

## Data Availability

All data produced in the present study are not available

